# Evaluating Observer Reliability and Diagnostic Accuracy of CT-LEFAT Criteria for Post-Treatment Head and Neck Lymphedema: A Prospective Blinded Comparative Analysis of Oncologist Human Inter-Rater Performance

**DOI:** 10.1101/2024.09.17.24313809

**Authors:** MD Anderson Head and Neck Cancer Symptom Working Group, Natalie A. West, Serageldin K. Attia, Zaphanlene Kaffey, Cem Dede, Samuel L. Mulder, Dina M. El-Habashy, Roger Neuberger, Mohamed A. Naser, Steven J. Frank, Shitong Mao, Holly McMillan, Brad Smith, David Rosenthal, Stephen Y. Lai, Katherine A. Hutcheson, Amy C. Moreno, Clifton D. Fuller

## Abstract

**Background:** Radiation-associated lymphedema and fibrosis (LEF) is a significant toxicity following radiation therapy (RT) for head and neck cancer (HNC) patients. Recently, the CT Lymphedema and Fibrosis Assessment Tool (CT-LEFAT) was developed to standardize LEF diagnosis through fat stranding visualized on CT. This study aims to evaluate the inter-observer reliability and diagnostic accuracy of the CT-LEFAT criteria.

**Materials and Methods:** This study retrospectively evaluated 26 HNC patients treated with RT that received a minimum of two contrast-enhanced CT scans. Qualitative review was conducted by five physician raters to assess the fat stranding observed on CT according to the CT-LEFAT criteria. Fleiss’ kappa analysis was used to assess the inter- and intra-rater reliability, and Receiver Operating Characteristic (ROC) Area Under the Curve (AUC) analysis was used to evaluate diagnostic accuracy.

**Results:** The inter-rater reliability across the six CT-LEFAT regions generally indicated a slight to fair agreement across all raters (0.04 ≤ kappa ≤ 0.36). Intra-observer agreement was generally fair to moderate (overall kappa=0.44). The ROC AUC analysis varied based on aggregation method used (0.60 ≤ average AUC ≤ 0.70).

**Conclusion:** This specific use-case evaluating CT-LEFAT criteria displays limited performance. This suggests that additional materials, such as further training, refinement of imaging methods, or other processes may be required before achieving clinically-ready diagnostic performance of LEF diagnosis.

## Introduction

Radiation-associated lymphedema and fibrosis (LEF) affects more than 90% of head and neck cancer (HNC) patients treated with radiation therapy^1,2^, which may lead to symptoms including swelling, pain, and limitation in range of motion that may decrease functionality and quality of life. Lymphedema is characterized by the abnormal accumulation of lymph fluid in tissues, presenting as swelling, while fibrosis involves thickening and hardening of tissue due to excess connective tissue formation. These conditions often occur as a progressive continuum or concurrently, particularly in HNC patients treated with surgery and/or radiation therapy (RT). Among those diagnosed, approximately 10% experience exclusively external LEF, affecting the face, submental region, and neck, while 40% experience only internal LEF, affecting the oral cavity, pharynx, and larynx. Additionally, around 50% of LEF patients experience both internal and external LEF^3^. Despite the prevalence of LEF and its impact, there is a notable gap in standardized methods for accurately evaluating these conditions^2,4,5^.

Traditionally, diagnosing LEF in post-cancer treatment patients has relied on clinical examinations, patient-reported symptom assessments, and various cutaneous surface measurement techniques^4^. Several scales are commonly used to evaluate LEF, each with distinct criteria and limitations. Overall, these scales tend to be subjective, relying on physician-based diagnosis, and do not address important parameters such as depth of LEF. The CTCAE Lymphedema and Fibrosis Scales (version 3.0), developed by the National Cancer Institute^6^, grade the severity of lymphedema and fibrosis but lack specificity for HNC patients and do not fully capture the functional impact. The ACS Lymphedema Scale, created by the American Cancer Society^7^, focuses on local and general affected areas but also misses detailed fibrosis assessment and HNC specificity. The Földi Scale^8^ grades both lymphedema and fibrosis severity comprehensively but is primarily used for limb lymphedema, making it less applicable to HNC. The MD Anderson Head and Neck Lymphedema (HNL) adaptation of the Földi Scale^9^ is a non-depth specific, global evaluation of all HNC regions and relies on subjective palpation, leading to variability between clinicians. The Patterson Scale^10^ focuses on endoscopic judgment of internal edema with limitations related to poor agreement in structures outside the supraglottis despite updates to the operating guidelines^17^. Finally, the initial Head and Neck External Lymphedema and Fibrosis (HN-ELAF) Assessment Criteria is a grading scale that used visible tissue edema, pitting, elevation effects, and fibrosis in its revised 5-step scale^10^.

To address the limitations of clinical and patient-reported assessments, the CT Lymphedema and Fibrosis Assessment Tool (CT-LEFAT)^2^ was developed as an imaging solution for LEF measurement. CT-LEFAT aims to provide an objective, imaging-based assessment that reduces subjectivity and offers a standardized, comprehensive method for evaluating LEF in HNC patients. It grades fat stranding and tissue changes at specific anatomical sites using CT scans, enhancing diagnostic accuracy and reproducibility. While CT-LEFAT shows major promise in the development of a standardized measurement tool, no variability and reliability findings were established with the initial study. We selected CT-LEFAT because it is a comprehensive tool that addresses the shortcomings of existing scales.

This study’s specific aim is to evaluate the inter-observer performance, namely, reliability and diagnostic accuracy, of the previously developed CT-LEFAT criteria, as applied to non-radiologist physician image examination. The objectives focus on prospective characterization of oncologist human rater assessment using standardized metrics in a cohort of HNC patients, along with analyzing inter-and intra-rater variability.

## Materials and Methods

In this prospective human-performance study, we implemented formal reporting guidance as per Enhancing the QUAlity and Transparency Of health Research Network guidance, using the *Guidelines for Reporting Reliability and Agreement Studies* (GRRAS)^12^.

### Patients

A convenience sample of 26 patients diagnosed with oropharyngeal cancer and treated with curative-intent RT at MD Anderson Cancer Center were retrospectively enrolled. The subset was sequentially sampled from an internal database of 104 patients after institutional review board (IRB) approval. The sample was limited to 26 sequentially sampled patients due to the large amount of time required for the review process. Patients included were treated with RT, had at least one diagnostic pre-RT CECT, and had at least one follow-up CECT within 3 months of lymphedema evaluation. The minimum time of follow-up CECT was 3-6 months following RT. Patients were treated with RT from 2017-2019 and contrast-enhanced computed tomography (CECT) scans were acquired from 2017-2020. Given the study’s anonymized design, the need for informed consent was waived. The medical records of the patients were retrieved and screened for demographic information, tumor characteristics, and lymphedema evaluation details and outcomes. Patient electronic health records were reviewed to establish the presence of lymphedema corresponding to each imaging study. For this study purposes, we used the MD Anderson Cancer Center Head and Neck Lymphedema staging^9^ as the clinical reference, with patients classified with a stage of 1a or higher considered to have lymphedema.

### CT imaging protocol

In this study, a minimum of one pre-radiotherapy (RT) scan per patient was included in addition to up to two follow-up scans. All CECT images were performed as part of the standard of care in accordance with the institutional protocol.

All CECT scans were acquired using standard-of-care imaging practice to reflect real-world implementation and were extracted from the MD Anderson Picture Archiving and Communication System (PACS) (Stentor/Intellispace Radiology 4.7, Philips Healthcare Informatics, Foster, CA, USA). Acquisition was performed using multidetector 64-row CT scanners (Discovery CT750 HD or Revolution HD CT scanners, GE HealthCare, Waukesha, WI, USA). The imaging parameters were as follows: revolution time of 0.60–1.00 seconds, pitch factor of 0.97, table feed of 19.38 mm per rotation, voltage of 120–140 kVp, an X-ray tube current of 159–640 mA (median 290), and slice thickness of 1-3 mm (median slice thickness reconstruction of 1mm across all cases). Images were acquired at a resolution of 512 × 512 pixels following a 90 second delay post-intravenous contrast administration.

### Imaging Data Preparation

The selected CECT images were retrieved from the PACS system and transferred to RayStation Treatment Planning System Version 11B (RaySearch, Stockholm, Sweden). To assess intra-observer agreement, scans from five subjects were duplicated five times, resulting in 25 repeats. In total, 129 individual images were reviewed. All images were anonymized and shuffled to ensure blinded evaluation. The window level was standardized to 350 and 40 HU for width and center levels, respectively.

A total of five physician raters were tasked with evaluating fat stranding^13,14^ using the CT-LEFAT protocol^2^, which involves a detailed grading system outlined as follows: fat stranding is graded on a 0–2 scale, with an option of N/A, across five specific locations at the level of the superior thyroid cartilage (axial reconstruction images). The facilitator scrolled through a region of slices near the level of the superior thyroid cartilage to allow the observers to reach consensus on the slice to use based on clarity of fat stranding. The grading criteria and methodology are detailed in Figure 1. To complete the evaluation of all cases, three sessions were conducted, during which raters independently submitted their assessments using REDCap electronic data capture tools hosted at MD Anderson Cancer Center^18,19^. REDCap (Research Electronic Data Capture) is a secure, web-based software platform designed to support data capture for research studies, providing 1) an intuitive interface for validated data capture; 2) audit trails for tracking data manipulation and export procedures; 3) automated export procedures for seamless data downloads to common statistical packages; and 4) procedures for data integration and interoperability with external sources. This instrument provided both descriptions and visual references for the regions of interest and included links to the published CT-LEFAT criteria^2^ to standardize training across all observers. All observers were blinded with regards to clincal findings or relevant diagnoses, and to other observer’s ratings. All readers were residency-trained clinicians with variable years of experience in radiation oncology. Readers included two radiation oncologists with over 10 years of experience, one radiation oncologist with 5 years of experience, and two trained radiation oncology postdoctoral fellows. They were blinded to both the image selection process and the lymphedema outcomes.

**Figure 1.**
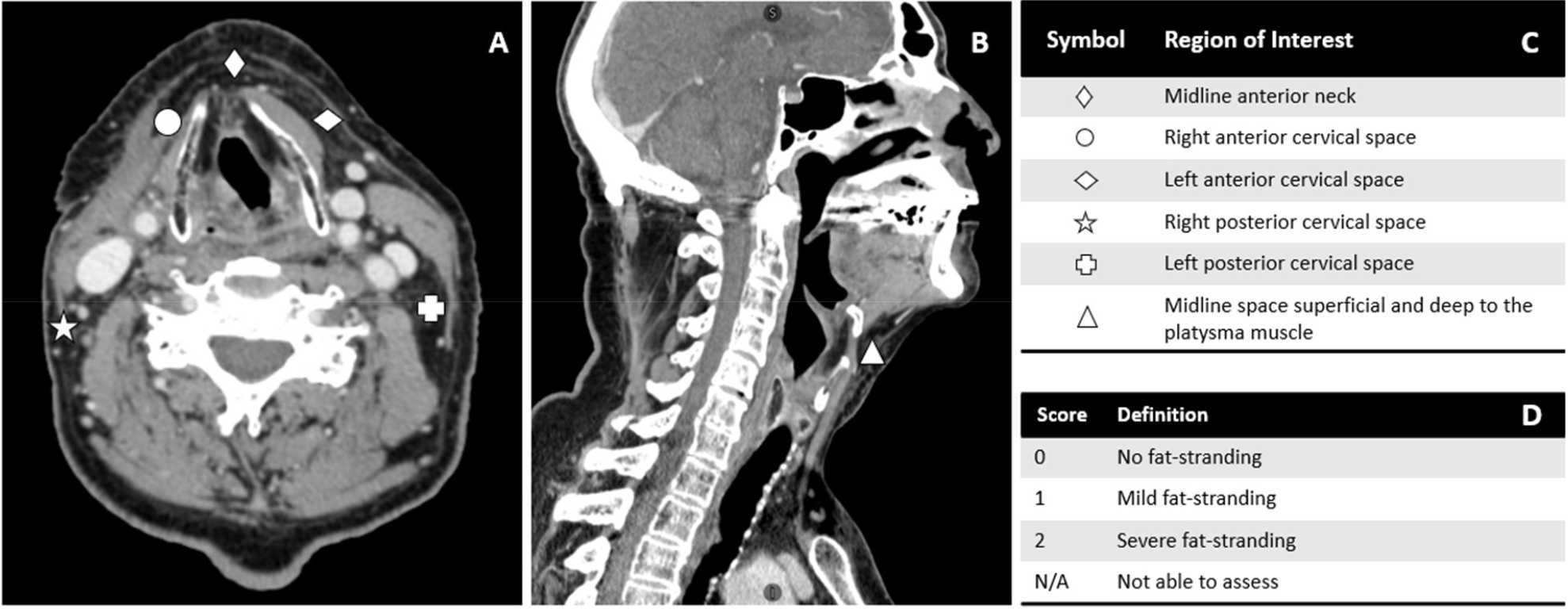
Grading criteria according to CT-LEFAT procedure. (A) shows five locations in the axial plane at the level of the superior thyroid cartilage. (B) displays the sixth grading region in the sagittal plane. (C) defines each region of interest and (D) defines scoring criterion. Adapted from Aulino et al.

### Statistical Analyses

Kappa values, following Fleiss’ method for multiple raters were obtained to establish the inter-rater and intra-rater reliability. The strength of agreement was evaluated using the following levels: zero as ‘poor’, 0.0-0.2 as ‘slight’, 0.021-0.4 as ‘fair’, 0.41-60 as ‘moderate’, 0.61-0.8 as ‘substantial’, and 0.81-1.0 as ‘almost perfect’^20^. To evaluate the validity of the CT-LEFAT criteria, the Receiver Operating Characteristic (ROC) Area Under the Curve (AUC) was calculated using different approaches to aggregate grading of fat stranding: 1) a combined score, calculated as the sum of grades across all six ROIs for each case, 2) maximum rule decision, where the highest score among the six ROIs was used as the representative index and 3) binarizing values of the CT-LEFAT grades to 0 for absent and 1 for moderate and severe fat stranding to produce and average binary index where values equal to or greater than 0.5 was indicative of lymphedema^2^.

**Figure 2.**
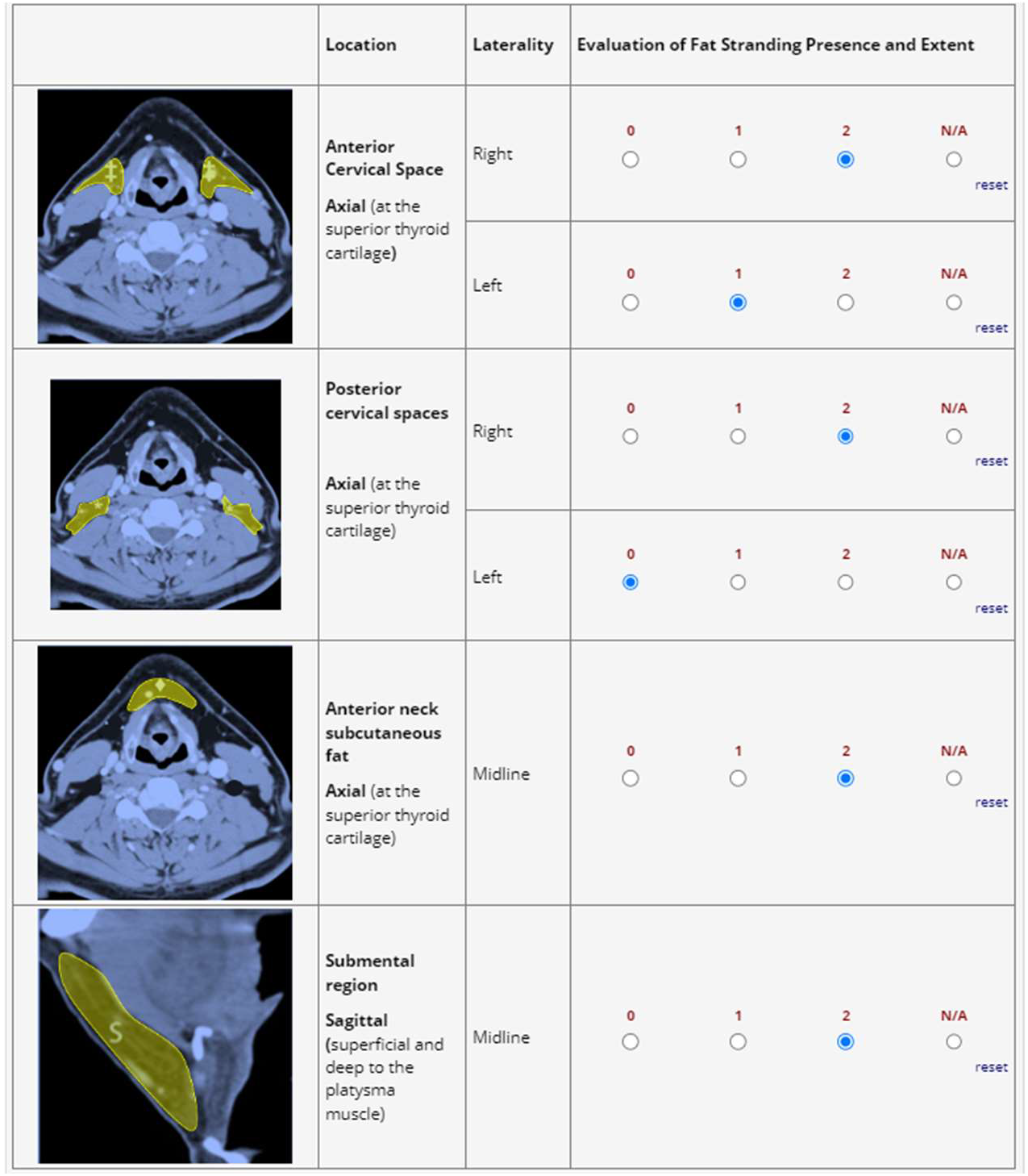
Representation of the REDCap survey tool used by raters for evaluation. ROIs delineated in the REDCap survey tool were not delineated during the review—raters had access to the example throughout the review.

## Results

The inter-rater reliability across the six CT-LEFAT regions generally indicated a slight to fair agreement across all raters with kappa levels ranging between 0.04-0.36 (median kappa=0.16). The anterior and posterior cervical regions exhibited slight agreement while the submental and anterior neck subcutaneous fat regions had fair agreement levels (Table 1).

**Table 1.** Inter-Rater Reliability Across ROIs for Different Time Points.

|  | **Baseline**<br>**(k)** | **Follow-up**<br>**1 (k)** | **Follow-up**<br>**2 (k)** | **Mean**<br>**(k)** | **Interpretation** |
| --- | --- | --- | --- | --- | --- |
| **Anterior Cervical - Left** | 0.12 | 0.12 | 0.22 | 0.15 | Slight agreement |
| **Anterior Cervical - Right** | 0.17 | 0.08 | 0.16 | 0.14 | Slight agreement |
| **Anterior Neck Subcutaneous Fat** | 0.24 | 0.22 | 0.21 | 0.22 | Fair agreement |
| **Posterior Cervical - Left** | 0.04 | 0.10 | 0.14 | 0.09 | Slight agreement |
| **Posterior Cervical - Right** | 0.09 | 0.11 | 0.12 | 0.11 | Slight agreement |
| **Submental Region (Sagittal)** | 0.33 | 0.25 | 0.36 | 0.31 | Fair agreement |

k: exact Fleiss kappa statistic

Intra-observer agreement was generally fair to moderate (overall kappa=0.44). The anterior cervical, anterior neck subcutaneous fat, and submental regions had stronger agreement compared to the posterior cervical regions (Table 2).

**Table 2.** Intra-Rater Agreement Across different ROIs and Raters.

|  | **Kappa** | **Interpretation** |
| --- | --- | --- |
| **Region** |  |  |
| Anterior Cervical - Left | 0.43 | Moderate agreement |
| Anterior Cervical - Right | 0.57 | Moderate agreement |
| Anterior Neck Subcutaneous Fat | 0.49 | Moderate agreement |
| Posterior Cervical - Left | 0.30 | Fair agreement |
| Posterior Cervical - Right | 0.31 | Fair agreement |
| Submental Region (Sagittal) | 0.55 | Moderate agreement |
| **Rater** |  |  |
| Rater A | 0.44 | Moderate agreement |
| Rater B | 0.62 | Substantial agreement |
| Rater C | 0.51 | Moderate agreement |
| Rater D | 0.39 | Fair agreement |
| Rater E | 0.24 | Fair agreement |

In the validation of the CT-LEFAT criteria for detecting lymphedema, the ROC AUC analysis showed different levels of diagnostic accuracy for the different aggregation methods. For the combined CT-LEFAT score, the mean AUC was 0.70. In contrast, when at least one non-zero score for any ROI was considered indicative of lymphedema (maximum rule), the AUC was lower, averaging 0.60 which was similar to the third approach where CT-LEFAT grades were binarized, demonstrating an average AUC of 0.63 (Figure 3).

**Figure 3.**
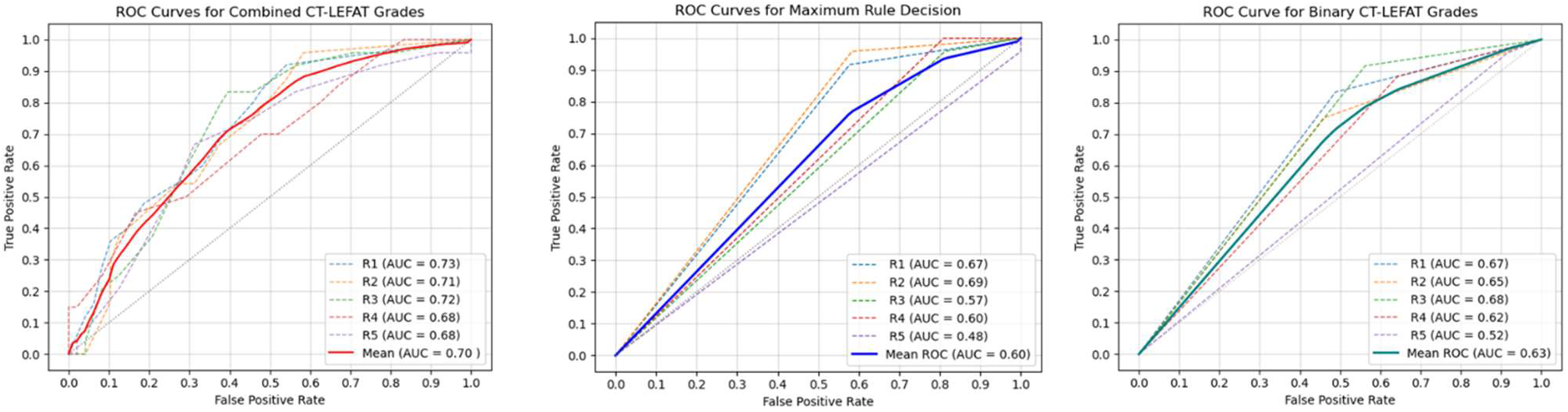
Receiver Operating Characteristic (ROC) curves for the CT-LEFAT Diagnostic Performance by individual raters (dashed lines) and average consensus (solid line): A) sum of CT-LEFAT grades B) Maximum rule decision of highest grade across all ROIs, and C) Averaged binary values across all ROIs.

## Discussion

Many radiotherapy patients will exhibit clinical lymphedema/fibrosis at some point in therapy, and as such there is an urgent need for effective severity grading and assessment of this prevalent iatrogenic disease state. However, the current standard of care for assessment of lymphedema/fibrosis requires manual grading of external lymphedema, surface measurement by a Certified Lymphedema Therapist and a separate internal lymphedema assessment by a Speech Language Pathologist or Otolaryngologist using the revised Patterson scale. These procedures require specialized expertise, and additional devices and testing, precluding ready integration into standard oncologic surveillance. Radiographic assessment of lymphedema/fibrosis would be exceptionally useful if validated, as surveillance imaging for oncologic surveillance using CT or MRI might be repurposed for assessment of lymphedema/fibrosis. Moreover, semi-quantitative objective measures of fibrosis provide a potential surrogate for pharmacologic interventions designed to ameliorate acute-to-chronic development or reversal of LEF. Ideally, said radiographic criteria would be reproducible in terms of inter-observer performance, localize disease severity spatially within the head and neck region, and provide distinct severity gradations for disease stratification. CT-LEFAT thus presents an enticing conceptual improvement in diagnosing and monitoring lymphedema. Despite this potential, human performance seems to be a limiting factor regarding non-radiologist utilization of the CT-LEFAT criteria in the observed study.

Our results show that this tool has low inter- and intra-reader agreement when measuring lymphedema, and this variance was amplified when stratified by region. This variance suggests that human performance is lacking in the diagnosis of lymphedema and fibrosis, demonstrating a marked need for deep learning in LEF. As current LEF assessment scales are harder to standardize^3^, one of the reasons for assessing this was to determine whether we can automate this process for machine learning-enabled interventions. However, given the observed human performance, it is unlikely that it will have high discriminate power in the absence of some other standard. This study identifies a marked need for standardized scales for LEF diagnosis; this can potentially be accomplished by leveraging imaging methods, evaluating LEF by site, and developing a physician training standard of LEF diagnosis.

### Comparison of Results to Existing Literature and Previous Studies

To our knowledge, this is the first independent study to assess the inter-reader reliability and intra-reader variability of CT-LEFAT as a radiographic assessment tool.

### Limitations and Potential Sources of Bias

As we undertook the proposed study as a prospective blinded study of human observer performance among oncologists and thus used previously acquired radiographic images, the images did not always correlate with time of diagnosis, which may lead to inaccurate LEF assessment. A prospective trial could improve on this by ensuring CT scans are obtained at time of diagnosis. LEF clinical diagnoses can be biased based off baseline imaging, and it is difficult to distinguish fat stranding from post-surgical or tumor effects on imaging alone^15^. There is also a possibility that prior fat stranding can influence diagnosis despite baseline images being obtained. This study included a baseline image in addition to temporal serial images for each patient to reflect clinical standards.

The MD Anderson Head and Neck Lymphedema (HNL) adaptation of the Földi Scale^9^ provides a global score of LEF. This presents a potential weakness in establishing a ground truth of LEF diagnosis, as global evaluation may misdiagnose LEF based on non-uniform distribution of clinical presentation of symptoms.

Additionally, this study only used five readers, those of which lacked formal radiology training, which could have affected the diagnostic accuracy when using this imaging-based tool. These results could be improved with a larger reader pool of those with formal radiology training. This configuration was inconsistent with the developers of CT-LEFAT, which included radiologists^2^. Knowing this, these results show that the standards of diagnosing LEF may vary across specialty, which needs to be investigated further.

### Future Research Directions

The quantitative potential of CT and MRI in the diagnosis and assessment of LEF is significant^15,16^. MRI stands out for its superior soft tissue contrast, enabling the differentiation between fluid accumulation and fibrotic changes without ionizing radiation exposure. With this, MRI shows promise in improved visualization as a radiographic tool to diagnose LEF.

Due to the diverse number of LEF assessment scales, there is a need to establish a radiological endpoint to diagnose LEF. The development of radiological tools, particularly with the potential of MRI, can optimize and expedite a move to deep learning as a discriminator in LEF, which can standardize LEF diagnosis. Through developing radiographic tools and standardizing LEF evaluation, we can determine if LEF is identifiable earlier through radiographic findings or clinical presentation, further improving LEF diagnosis.

## Conclusion

CT-LEFAT showed limited performance in a radiation oncologist-specific human performance test. However, this suggests that either additional curricular training, further refinement of instructional materials, additional imaging methods, or other augmentation may be required to achieve clinically-ready performance as a diagnostic discriminator of lymphedema severity.

## Supporting information

Supplemental Figure 1: Presentation used as training prior to physician's review. Raters were also provided access to the reference for supplement.

Supplemental Table 1: REDCap Data Dictionary Codebook used to evaluate fat stranding.

## Data Availability

The anonymized dataset is available in Figshare (10.6084/m9.figshare.27040849).

## References

1. Ridner, S. H., Dietrich, M. S., Niermann, K., Cmelak, A., Mannion, K., & Murphy, B. (2016). A Prospective Study of the Lymphedema and Fibrosis Continuum in Patients with Head and Neck Cancer. Lymphatic research and biology, 14(4), 198–205. 10.1089/lrb.2016.0001

2. Aulino JM, Wulff-Burchfield EM, Dietrich MS, Ridner SH, Niermann KJ, Deng J, Rhoten BA, Doersam JK, Jarrett LA, Mannion K, Murphy BA. Evaluation of CT Changes in the Head and Neck After Cancer Treatment: Development of a Measurement Tool. Lymphat Res Biol. 2018 Feb;16(1):69–74. doi: 10.1089/lrb.2017.0024. PMID: 29432066; PMCID: PMC5810430.

3. Deng J, Ridner SH, Dietrich MS, Wells N, Wallston KA, Sinard RJ, Cmelak AJ, Murphy BA. Prevalence of secondary lymphedema in patients with head and neck cancer. J Pain Symptom Manage. 2012 Feb;43(2):244–52. doi: 10.1016/j.jpainsymman.2011.03.019. Epub 2011 Jul 30. PMID: 21802897.

4. Arends CR, Lindhout JE, van der Molen L, Wilthagen EA, van den Brekel MWM, Stuiver MM. A systematic review of validated assessments methods for head and neck lymphedema. Eur Arch Otorhinolaryngol. 2023 Jun;280(6):2653–2661. doi: 10.1007/s00405-023-07841-0. Epub 2023 Feb 10. PMID: 36763153; PMCID: PMC10175329.

5. Deng J, Ridner SH, Dietrich MS, Wells N, Murphy BA. Assessment of external lymphedema in patients with head and neck cancer: a comparison of four scales. Oncol Nurs Forum. 2013 Sep;40(5):501–6. doi: 10.1188/13.ONF.501-506. PMID: 23989023.

6. Cancer Therapy Evaluation Program, Common Terminology Criteria for Adverse Events, Version 3.0, DCTD, NCI, NIH, DHHS. March 31, 2003 (http://ctep.cancer.gov), Publish Date: August 9, 2006.

7. The American Cancer Society Medical and Editorial Content Team. “Lymphedema.” American Cancer Society, www.cancer.org/cancer/managing-cancer/side-effects/swelling/lymphedema.html. Accessed 11 July 2024.

8. Földi M, Földi E. Foldi’s Textbook of Lymphology. 3rd ed. München: Urban & Fischer; 2012.

9. Smith BG, Lewin JS. Lymphedema management in head and neck cancer. Curr Opin Otolaryngol Head Neck Surg. 2010 Jun;18(3):153–8. doi: 10.1097/MOO.0b013e32833aac21. PMID: 20463478; PMCID: PMC4111092.

10. Patterson JM, Hildreth A, Wilson JA. Measuring edema in irradiated head and neck cancer patients. Ann Otol Rhinol Laryngol. 2007; 116(8): 559–564.

11. Deng, J., Ridner, S. H., Wells, N., Dietrich, M. S., & Murphy, B. A. (2015). Development and preliminary testing of head and neck cancer related external lymphedema and fibrosis assessment criteria. European journal of oncology nursing : the official journal of European Oncology Nursing Society, 19(1), 75–80. 10.1016/j.ejon.2014.07.006

12. Kottner J, Audigé L, Brorson S, Donner A, Gajewski BJ, Hróbjartsson A, Roberts C, Shoukri M, Streiner DL. Guidelines for Reporting Reliability and Agreement Studies (GRRAS) were proposed. J Clin Epidemiol. 2011 Jan;64(1):96–106. doi: 10.1016/j.jclinepi.2010.03.002. Epub 2010 Jun 17. PMID: 21130355.

13. Filippone A, Cianci R, Pizzi AD, Esposito G, Pulsone P, Tavoletta A, et al. CT findings in acute peritonitis: A pattern-based approach. Diagnostic Interv Radiol. 2015;21:435–40.

14. Thornton E, Mendiratta-Lala M, Siewert B, Eisenberg RL. Patterns of fat stranding. AJR Am J Roentgenol. 2011 Jul;197(1):W1–14. doi: 10.2214/AJR.10.4375. PMID: 21700969.

15. Bronstein AD, Nyberg DA, Schwartz AN, Shuman WP, Griffin BR. Soft-tissue changes after head and neck radiation: CT findings. AJNR. 1989;10:171–175.

16. Shin SU, Lee W, Park EA, Shin CI, Chung JW, Park JH. Comparison of characteristic CT findings of lymphedema, cellulitis, and generalized edema in lower leg swelling. Int J Cardiovasc Imaging 2013; 29:135–143.

17. Starmer H, Cherry MG, Patterson J, Young B, Fleming J. Assessment of Measures of Head and Neck Lymphedema Following Head and Neck Cancer Treatment: A Systematic Review. Lymphat Res Biol. 2023 Feb;21(1):42–51. doi: 10.1089/lrb.2021.0100. Epub 2022 Jun 9. PMID: 35679595.

18. PA Harris, R Taylor, R Thielke, J Payne, N Gonzalez, JG. Conde, Research electronic data capture (REDCap) – A metadata-driven methodology and workflow process for providing translational research informatics support, J Biomed Inform. 2009 Apr;42(2):377–81.

19. PA Harris, R Taylor, BL Minor, V Elliott, M Fernandez, L O’Neal, L McLeod, G Delacqua, F Delacqua, J Kirby, SN Duda, REDCap Consortium, The REDCap consortium: Building an international community of software partners, J Biomed Inform. 2019 May 9 [doi: 10.1016/j.jbi.2019.103208]

20. Landis JR, Koch GG. The measurement of observer agreement for categorical data. Biometrics. 1977 Mar;33(1):159-74. PMID: 843571.

