## Supplemental Figure 1: Presentation used as training prior to physician's review. Raters were also provided access to the reference for supplement. for "Evaluating Observer Reliability and Diagnostic Accuracy of CT-LEFAT Criteria for Post-Treatment Head and Neck Lymphedema: A Prospective Blinded Comparative Analysis of Oncologist Human Inter-Rater Performance"

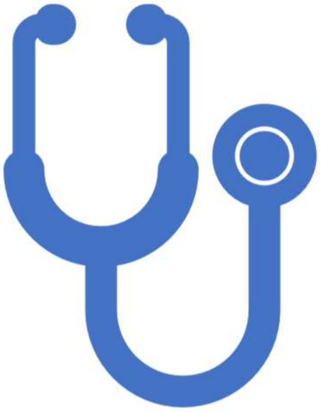

09/07/2023

### CT-LEFAT Physician's Review Tutorial

1

#### CT-LEFAT Background

- RT can lead to development of late toxicities like lymphedema and fibrosis (LEF) in H+N patients
- Measurement and documentation of LEF is challenging due to anatomic complexities of H+N
- Radiologic review conventionally has not considered soft tissue damage and has no standardized method of grading soft tissue changes post RT

Physician's Review

2

2

### CT-LEFAT grades fat stranding

- Grades fat stranding in 6 locations on a scale of 0 to 2
- Fat stranding- abnormal increased attenuation in fat
- Can be seen in inflammatory, infections, or malignant conditions
- Fat stranding may identify patients with LEF

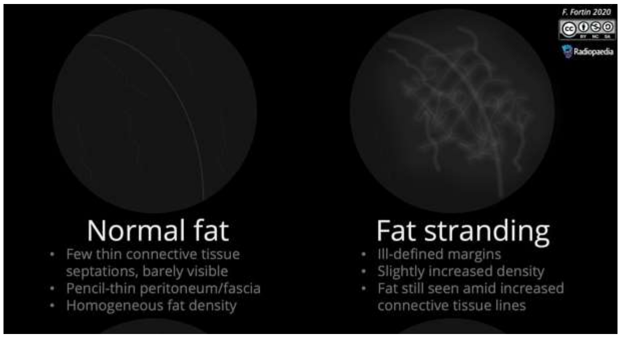


https://radiopaedia.org/articles/fat-stranding-summary-3



### 6 locations of fat stranding grading

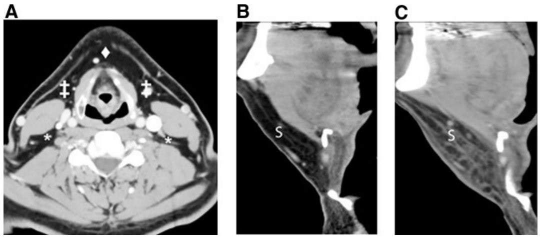


[Aulino et al]

- bilateral anterior cervical space
- posterior cervical space
- midline anterior neck subcutaneous fat at the level of the superior thyroid cartilage as visible on axial reconstructed images and within the submental region on sagittal reformatted images
- Window width 350 HU, window level 40 HU

### Measure AP diameter of epiglottis

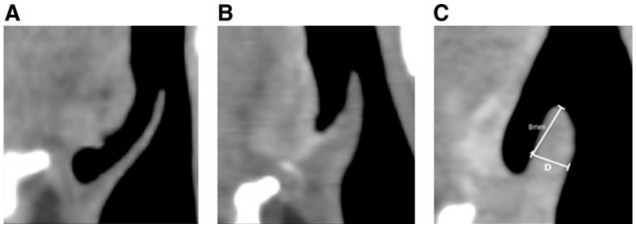


- Epiglottic submucosal edema causes a volume change and is common post RT
- Sagittal reformatted images are used to examine volume change
- AP diameter of epiglottis is measured

Physician's Review 5

[Aulino et al]

5

### Measure Prevertebral Soft Tissue Thickness (PVST)

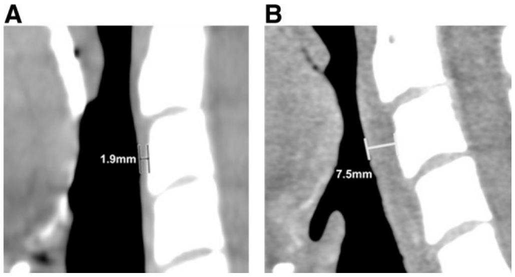


[Aulino et al]

- PVST can increase post RT
- A shows the baseline measurement before radiation
- B shows the thickened prevertebral soft tissue 8 months post RT

Physician's Review 6

6

### CT-LEFAT Protocol

1. Score 6 specified locations for fat stranding grading
2. Measure AP diameter of epiglottis
3. Measure PVST
4. Record scores and measurements in REDCap
