## Supplemental Table 1: REDCap Data Dictionary Codebook used to evaluate fat stranding. for "Evaluating Observer Reliability and Diagnostic Accuracy of CT-LEFAT Criteria for Post-Treatment Head and Neck Lymphedema: A Prospective Blinded Comparative Analysis of Oncologist Human Inter-Rater Performance"

| # | Variable / Field Name | Field Label<br>*Field Note* | Field Attributes (Field Type, Validation, Choices, Calculations, etc.) |
|---|---|---|---|
| **Instrument: Subject ID (subject_id)** | | | |
| 1 | [subject_id] | Subject ID | text, Identifier |
| 2 | [subject_last_name] | Last Name | text |
| 3 | [subject_id_complete] | Section Header: *Form Status*<br>Complete? | dropdown<br>0 Incomplete<br>1 Unverified<br>2 Complete |
| **Instrument: Reader (reader)** | | | |
| 4 | [description] | The objective of this form is to validate the evaluation criteria developed for assessing radiation-induced changes (CT-LEFAT) as outlined in the original publication by Aulino et al | descriptive |
| 5 | [img_modality_lefat] | Please indicate the modality used for the assessment<br>*Please select image modality/protocol you used for the evaluation* | dropdown<br>1 CECT (W/L 350/40 HU)<br>2 MRI T2 FS<br>3 MRI T1+/-C |
| 6 | [study_date_lefat] | Imaging Study Date<br>*Please enter study date YYYY-MM-DD* | text (date_ymd)<br>Field Annotation: @HIDEBUTTON |
| 7 | [reader_name_lefat] | Reader's Name<br>*Please enter your first and last name. You are logged in as [user-fullname]* | text, Required, Identifier<br>Field Annotation: @DEFAULT="[user-fullname]" |
| 8 | [reader_initials_lefat] | Reader's Initials | text, Required, Identifier<br>Field Annotation: @USERNAME |
| 9 | [description_fat_stranding] | Fat Stranding Evaluation:Please evaluate and grade the presence and extent of fat stranding in the following locations on a scale of 0-2 where 0-normal, 1-mild changes, and 2-advanced. If a location is not evaluable, please select N/A. Location Laterality Evaluation of Fat Stranding Presence and Extent Anterior Cervical Space Axial (at the superior thyroid cartilage) Right {ant_cerv_r} Left {ant_cerv_l} Posterior cervical spaces Axial (at the superior thyroid cartilage) Right {post_cerv_r} Left {post_cerv_l} Anterior neck subcutaneous fat Axial (at the superior thyroid cartilage) Midline {ant_scf} Submental region Sagittal (superficial and deep to the platysma muscle) Midline {submental} | descriptive |
| 10 | [ant_cerv_r] | Anterior cervical space (Right) | radio (Matrix), Required<br>0 0<br>1 1<br>2 2<br>3 N/A |
| 11 | [ant_cerv_l] | Anterior cervical space (Left) | radio (Matrix), Required<br>0 0<br>1 1<br>2 2<br>3 N/A |
| 12 | [post_cerv_r] | Posterior cervical space (Right) | radio (Matrix), Required<br>0 0<br>1 1<br>2 2<br>3 N/A |
| 13 | [post_cerv_l] | Posterior cervical space (Left) | radio (Matrix), Required<br>0 0<br>1 1<br>2 2<br>3 N/A |
| 14 | [ant_scf] | Anterior neck subcutaneous fat (midline) | radio (Matrix), Required<br>0 0<br>1 1<br>2 2<br>3 N/A |
| 15 | [submental] | Midline sagittal submental region | radio (Matrix), Required<br>0 0<br>1 1<br>2 2<br>3 N/A |
| 16 | [submucosa_edema] | Submucosal edema evaluation: Please evaluate the volume changes using diameter of the following structures: AP diameter of the epiglottis, calculated 8 mm from the free margin (superior extent) to the anterior surface of the epiglottis, and then to the closest point on the posterior surface. Epiglottis Diameter (in mm): {epigl_thickness} Prevertebral soft tissue (PVST) at the mid-C3 level PVST Diameter (in mm): {pvst_thickness} | descriptive |
| 17 | [epigl_thickness] | Please enter the epiglottis thickness in millimeters | text (number)<br>Custom alignment: RH |
| 18 | [pvst_thickness] | Please enter the prevertebral soft tissue thickness (PVST) at the level of mid-C-3 in millimeters | text (number)<br>Custom alignment: RH |
| 19 | [eval_date_lefat] | Evaluation Date | text (date_ymd) |
